# Influence of social determinants of health on quality of life in patients with multimorbidity

**DOI:** 10.1101/2024.01.12.24301228

**Authors:** José María Ruiz-Baena, Aida Moreno-Juste, Beatriz Poblador-Plou, Marcos Castillo-Jimena, Amaia Calderón-Larrañaga, Cristina Lozano-Hernández, Antonio Gimeno-Miguel, Luis A Gimeno-Feliú, MULTIPAP GROUP

## Abstract

**Background:** Multimorbidity, especially among older patients, is one of the biggest challenges faced by modern medicine, and is influenced by social determinants of health, giving rise to health inequalities in the population. Here, we sought to determine the influence of social determinants of health on quality of life in patients with multimorbidity.

**Methods and Materials:** This cross-sectional observational study included 573 patients aged 65–74 with multimorbidity (≥3 diseases) and polypharmacy (≥5 drugs). Corresponding data was taken drawn from the Spanish MULTIPAP study, and included social and demographic variables, and data on health-related quality of life and overall self-perceived health status, assessed using the 5-level version of the EuroQol 5-dimensional questionnaire (EQ-5D-5L). Descriptive, bivariate and multivariate analyses with logistic regression models were performed.

**Results:** Mean patient age was 69.7 years, 55.8% of patients were female, 59.7% belonged to low social classes (V, VI), a monthly income of 1051–€1850 predominated, and the median number of diseases in the same patient was 6. Factors associated with higher quality of life were male gender (OR=1.599, p=0.013), a higher educational level (OR=1.991, p=0.036), an absence of urban vulnerability (OR=1.605, p=0.017), and the presence of medium social support (OR=1.689, p=0.017). Having a higher number of diseases was associated with poorer quality of life (OR=0.912, p=0.017).

**Conclusions:** Our findings describe associations between social determinants of health and quality of life in patients aged 65–74 years with multimorbidity and polypharmacy. More illnesses, female gender, a lower education level, urban vulnerability, and less social support are associated with poorer quality of life, underscoring the need for a biopsychosocial approach in patient care.

## Introduction

The last century has seen an unprecedented increase in human life expectancy. Inherent to this phenomenon is a parallel increase in the prevalence of chronic diseases for which old age is a key risk factor, including hypertension, arthrosis, and diabetes [1].

Multimorbidity is defined as the coexistence of several chronic diseases in the same patient [2], and affects an estimated 81.5% of individuals aged ≥85 years, 62% of those aged 65–74 years, and 50% of those aged >65 years [3]. Coexistence of several chronic diseases is a very common phenomenon, and has turned multimorbidity into one of the greatest challenges facing modern healthcare systems.

Some studies have demonstrated exponential increases in disability when the number of chronic diseases increases, and have characterized the effects of multimorbidity on health outcomes such as physical functioning, mental health, and quality of life [4]. Health-related quality of life (HRQoL) is a multi-dimensional concept used to examine the impact of health status on quality of life, and can be measured in several ways, both objective and subjective. It also includes the self-perception of the individual’s living situation, and their attitudes towards modifying and improving their conditions in relation to their goals, expectations, values and interests [5].

The mechanisms underlying multimorbidity are diverse and complex. On the one hand are biological factors inherently related to the aging process itself, including gender, cellular senescence, stem cell aging, and DNA methylation. On the other hand are factors such as lifestyle (e.g., diet, sedentarism, sleep quality, use of harmful substances like alcohol or other drugs), social and community-related variables (e.g., social support, loneliness, adverse childhood experiences), and work conditions, all of which are conditioned by cultural, environmental and socio-economic factors (urbanism, educational level) [6]. Although multimorbidity relates to the presence of diseases, it should not be viewed solely from a biomedical perspective, and requires a biopsychosocial, patient-focused approach. Primary care can play a central role in orchestrating a multifactorial approach to the management of pluri-pathology, with the goal of maximizing patient quality of life and minimizing potential morbidity and disability [7]. Considering how social factors influence these variables is essential to help achieve this goal.

Social determinants of health (SDH) are defined as the conditions in which people are born, grow, work, live, and age, including a combination of socio-economic, environmental, community and work-related factors, apart from those related to healthcare systems. Historically, SDH have been one of the main sources of societal inequities [8].

A better understanding of the relationship between SDH and health outcomes in older patients with multimorbidity is essential to help improve quality of care of these patients. The aim of this study was to examine how SDH influence quality of life in primary care patients aged 65–74 with multimorbidity.

## Materials and Methods

### Design and setting of the study

We conducted a descriptive cross-sectional study using baseline information recorded for participants enrolled in the MULTIPAP cluster-randomized controlled trial (RCT), which sought to improve drug prescription in young-old adults with multimorbidity and polypharmacy through a complex intervention in primary care. A detailed description of the design, methods, and intervention of the MULTIPAP trial has been published elsewhere [9].

Briefly, the MULTIPAP RCT involved the voluntary participation of 117 family physicians who recruited a total of 573 patients from 38 Spanish primary care health centres in 3 different regions (Andalucía, Madrid, and Aragón) between November 8, 2016, and January 31, 2017 [10]. Patients were randomly included in the study if they were aged 65–74 years, had multimorbidity (defined as the presence of 3 or more chronic diseases) and polypharmacy (defined as taking 5 or more different drugs during the preceding 3 months), had visited their family doctor at least once in the preceding year, and were able to follow the study requirements. Institutionalized patients and those with a life expectancy of less than 12 months and/or with a disease that in their physician’s opinion would not allow them to follow the study requirements were excluded. The recruiting physicians provided the patient with detail information about the study, they confirmed the patient’s eligibility and obtained the patient’s written informed consent.

This study was approved by the Clinical Research Ethics Committees of Aragón (CEICA, PI15/0217) and was favourably evaluated by the Research Ethics Committee of the Province of Malaga and by the Central Committee of Primary Care Research of the Community of Madrid.

### Study variables

All data were obtained through face-to-face surveys during baseline patient consultations and recorded using an electronic case report form. The information recorded for each patient is indicated below.

Independent variables included for each patient were as follows: date of birth; gender; nationality; marital status; family composition; housing indicators; self-perceived disturbances in the neighbourhood; social class, divided into 6 categories from I (highest) to VI (lowest) according to the Spanish 2017 National Health Survey and then dichotomized into high (I through IV) and low (V and VI); educational level according to the Spanish 2011 Statistical Office’s Population and Housing Census questionnaire [11]; occupation according to the Spanish Classification of Occupations CNO–11 [12]; socioeconomic status assessed based on monthly net income per household, expressed as multiples of the minimum wage; self-perceived social support measured using the Spanish version of the Duke Unc-11 Functional Social Support (DUFSS) questionnaire[13]; and the number of chronic conditions registered in the patient’s electronic health records.

Health-related quality of life and overall self-perceived health status, measured using the 5-level version of the EuroQol 5-dimensional questionnaire (EQ-5D-5L), was assessed as a dependent variable. The EQ-5D-5L considers 5 dimensions (mobility, self-care, daily activities, pain/discomfort, and anxiety/depression), and requires patients to rate their level of impairment for each item (5 levels, from 1 [no problems] to 5 [being unable to do/having extreme problems]). According to the responses to each item, an overall index score (ranging from 0–1) was calculated for each patient using the EQ-5D-5L Crosswalk Index Value Calculator. This calculator is available on the EuroQol website and provides validated results for the Spanish population obtained from a matrix of transition probabilities between health states [14].

### Statistical analysis

We described the characteristics of the study population as frequencies and/or the mean and standard deviation (sd). Subsequently, we analysed the distribution of patient quality of life according to the study variables. The Kolmogorov-Smirnov test indicated that quality of life followed a non-normal distribution. Therefore, the Mann Whitney U-test and the Kruskal-Wallis test were used for comparisons between 2 and >2 groups, respectively. In the case of the Kruskal-Wallis test, the Dwass-Steel-Critchlow-Fligner post-hoc test was performed for all variables for which statistical significance was found, to determine the specific pairs of variables for which the differences were significant. Results were expressed as the median and corresponding interquartile range (IQR).

Finally, we conducted a multivariate analysis to test the association between independent study variables and patient quality of life, using a logistic regression model for the EQ-5D-5L scale. To this end, the EQ-5D-5L variable was transformed into a dichotomised variable with 2 possible outcomes (low/high), depending on whether the patient’s score was below or above the median, respectively. This categorization was performed to obtain homogeneous groups.

All statistical analyses were conducted in STATA software (Version 17.0, StataCorp LLC, College Station, TX, USA) and Jamovi (Version 2.3.5) [17].

## Results

The main sociodemographic characteristics of the study population are shown in Table 1. Mean age was 69.7 years, and 55.8% of patients were women. A large portion of the sample (59.7%) corresponded to low social classes (V, VI). Monthly income was classified as medium (defined as a monthly salary of €1051–€1850) in the largest proportion of patients (42.6%). Finally, the median number of diseases in the same patient was 6 (IQR 4–7).

**Table 1.** Main sociodemographic characteristics of the study population.

| n | 573 |
| --- | --- |
| Age (mean (SD)) | 69.69 (2.68) |
| Gender = Female (%) | 320 (55.8) |
| Nationality = Spanish (%) | 564 (98.4) |
| Marital status (%) |  |
| Married | 433 (75.6) |
| Separated | 27 (4.7) |
| Single | 23 (4.0) |
| Widower | 90 (15.7) |
| Educational level (%) |  |
| Did not complete primary studies | 264 (46.1) |
| Completed primary studies | 235 (41.0) |
| Degree or higher | 74 (12.9) |
| Occupation (%) |  |
| Elementary occupations | 223 (38.9) |
| Essential sectors | 241 (42.1) |
| Technicians, office employment | 109 (19.0) |
| Social class (%) |  |
| I - Professional occupations | 35 (6.1) |
| II - Managerial and technical occupations | 26 (4.5) |
| III - Skilled non-manual occupations | 75 (13.1) |
| IV - Skilled manual occupations | 95 (16.6) |
| V - Partly skilled manual occupations | 205 (35.8) |
| VI - Unskilled manual occupations | 137 (23.9) |
| Monthly income (%) |  |
| Low - ≤€1050 | 163 (28.4) |
| Medium - €1051–€1850 | 244 (42.6) |
| High - ≥€1851 | 145 (25.3) |
| Unknown | 21 (3.7) |
| Home size (m<sup>2</sup>), mean (SD) | 94.03 (43.39) |
| Lives alone = Yes (%) | 101 (17.6) |
| Number of cohabitants, mean (SD) | 2.30 (0.68) |
| Urban vulnerability<sup>1</sup> = Yes (%) | 203 (35.6) |
| EQ-5D-5L, mean (SD) | 0.77 (0.20) |
| Functional social support, mean (SD) | 43.82 (8.75) |
| Number of diseases, median [IQR] | 6.00 [4.00, 7.00] |
| <sup>1</sup>Patients reported having problems with at least 4 of the 9 items assessed |  |

The results of a univariate analysis of the relationship between independent variables (analysed separately) and quality of life are shown in Table 2. Median quality of life was lower in women versus men; in patients from a low versus high social class; in patients in a situation of urban vulnerability; in patients with a low versus high monthly income; in patients with a lower educational level; and in patients with a lower level of social support.

**Table 2.** Quality of life (EQ-5D-5L) distribution according to social determinants of health: results of a univariate analysis.

| Variable | Median [IQR] | p-value |
| --- | --- | --- |
| **Gender** |  | <0.001 |
| Female | 0.790 [0.659–0.887] |  |
| Male | 0.857 [0.744–0.911] |  |
| **Social class** |  | 0.005 |
| Low (I–IV) | 0.799 [0.662–0.910] |  |
| High (V–VI) | 0.838 [0.737–0.924] |  |
| **Lives alone** |  | 0.244 |
| Yes | 0.800 [0.653–0.910] |  |
| No | 0.818 [0.694–0.910] |  |
| **Urban vulnerability<sup>1</sup>** |  | <0.001 |
| Yes | 0.769 [0.642–0.887] |  |
| No | 0.838 [0.711–0.910] |  |
| **Monthly income** |  | 0.004<sup>2</sup> |
| Low ≤€1050 | 0.799 [0.641–0.893] |  |
| Medium €1051–1850 | 0.818 [0.694–0.910] |  |
| High ≥€1851 | 0.846 [0.743–0.932] |  |
| **Educational level** |  | 0.005<sup>3</sup> |
| Did not complete primary studies | 0.818 [0.692–0.910] |  |
| Completed primary studies | 0.800 [0.662–0.910] |  |
| Degree or higher | 0.877 [0.781–0.932] |  |
| **Occupation** |  | 0.075 |
| Elementary occupations | 0.817 [0.662–0.910] |  |
| Essential sectors | 0.818 [0.694–0.910] |  |
| Technicians, office employments | 0.841 [0.723–0.932] |  |
| **Marital status** |  | 0.083 |
| Married | 0.818 [0.699–0.910] |  |
| Separated | 0.756 [0.649–0.910] |  |
| Single | 0.781 [0.682–0.921] |  |
| Widower | 0.809 [0.600–0.857] |  |
| **Social support** |  | <0.001<sup>4</sup> |
| Lower tertile | 0.784 [0.649–0.874] |  |
| Medium tertile | 0.857 [0.740–0.919] |  |
| Higher tertile | 0.818 [0.694–0.910] |  |

1. Patients reported having problems with at least 4 of the 9 items assessed
2. After applying the corresponding post-hoc pairwise comparison, significant differences were observed between the *Low* and *High* groups ( $p = 0.003$ )
3. After applying the corresponding post-hoc pairwise comparison, significant differences were observed between the groups *Did not complete primary studies* and *Degree or higher* ( $p = 0.003$ ), as well as *Completed primary studies* and *Degree or higher* ( $p = 0.01$ )
4. After applying the corresponding post-hoc pairwise comparison, significant differences were observed between the *Lower tertile* and *Medium tertile* groups ( $p < 0.001$ )

Table 3 shows the results of a multivariate analysis that examined the association between quality of life and several social variables. The analysis identified the following statistically significant risk factors for worse quality of life (odds ratio [OR], p-value): female sex (OR=1.599, p=0.013); lower versus higher educational level (OR=1.991, p=0.036); a situation of urban vulnerability (OR=1.605, p=0.017); and low versus medium level of social support (OR=1.689, p=0.017). Furthermore, a higher number of diseases was associated with worse quality of life (OR=0.912, p=0.017).

**Table 3.**
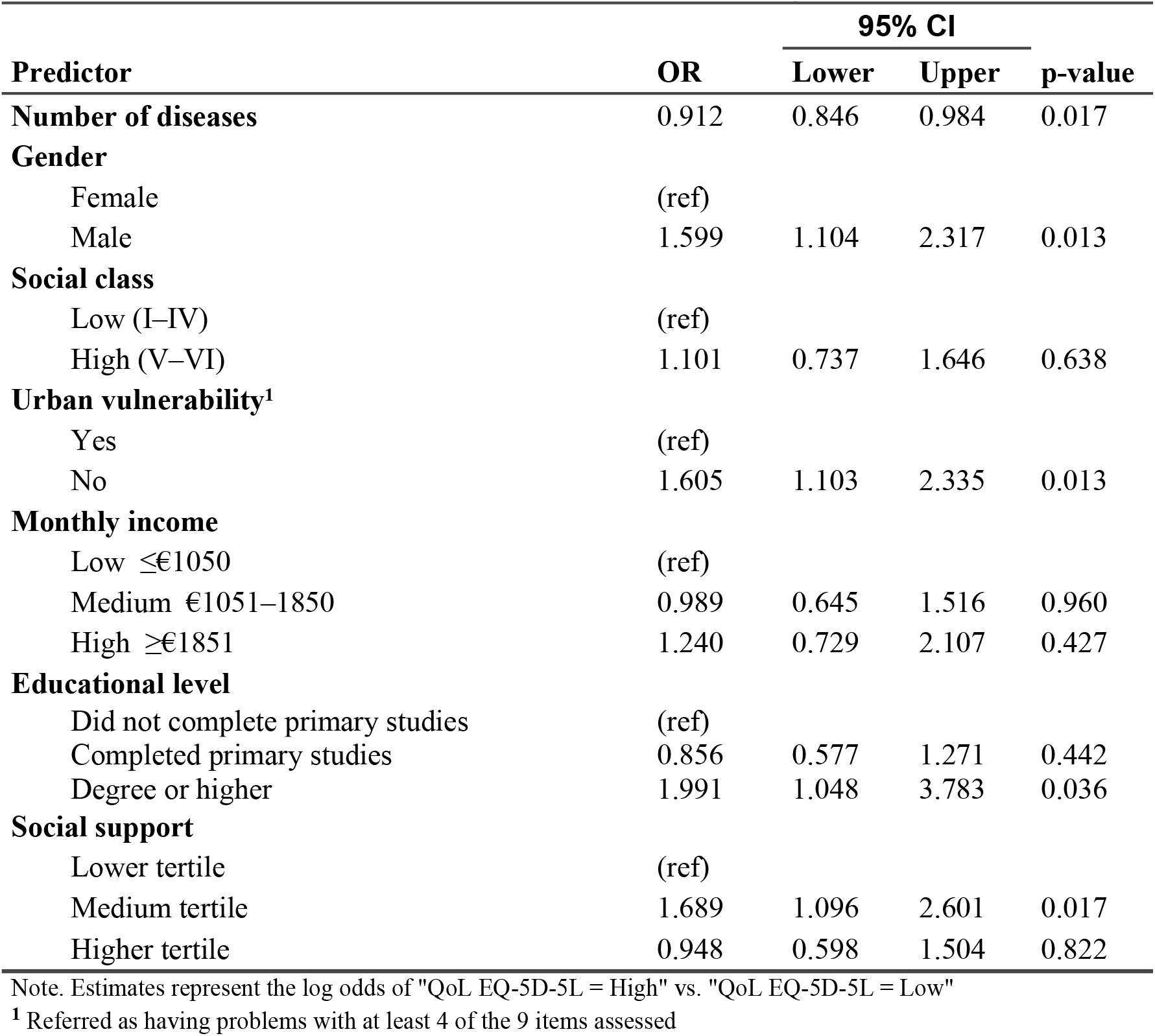
Association between quality of life (EQ-5D-5L) and variables related to social determinants of health (multivariate analysis)

| Predictor | OR | 95% CI |  | p-value |
| --- | --- | --- | --- | --- |
|  |  | Lower | Upper |  |
| **Number of diseases** | 0.912 | 0.846 | 0.984 | 0.017 |
| **Gender** |  |  |  |  |
| Female | (ref) |  |  |  |
| Male | 1.599 | 1.104 | 2.317 | 0.013 |
| **Social class** |  |  |  |  |
| Low (I–IV) | (ref) |  |  |  |
| High (V–VI) | 1.101 | 0.737 | 1.646 | 0.638 |
| **Urban vulnerability<sup>1</sup>** |  |  |  |  |
| Yes | (ref) |  |  |  |
| No | 1.605 | 1.103 | 2.335 | 0.013 |
| **Monthly income** |  |  |  |  |
| Low ≤€1050 | (ref) |  |  |  |
| Medium €1051–1850 | 0.989 | 0.645 | 1.516 | 0.960 |
| High ≥€1851 | 1.240 | 0.729 | 2.107 | 0.427 |
| **Educational level** |  |  |  |  |
| Did not complete primary studies | (ref) |  |  |  |
| Completed primary studies | 0.856 | 0.577 | 1.271 | 0.442 |
| Degree or higher | 1.991 | 1.048 | 3.783 | 0.036 |
| **Social support** |  |  |  |  |
| Lower tertile | (ref) |  |  |  |
| Medium tertile | 1.689 | 1.096 | 2.601 | 0.017 |
| Higher tertile | 0.948 | 0.598 | 1.504 | 0.822 |

Note. Estimates represent the log odds of "QoL EQ-5D-5L = High" vs. "QoL EQ-5D-5L = Low"
<sup>1</sup> Referred as having problems with at least 4 of the 9 items assessed

## Discussion

### Main findings

We observed significant associations between several of the SDH studied and quality of life in patients aged 65–74 with multimorbidity and polypharmacy. Specifically, the following independent variables were correlated with poorer quality of life: female gender; lower educational level; living in a neighbourhood in a situation of urban vulnerability; a lower level of social support; and a higher number of concomitant diseases.

### Comparison with other studies

While the literature highlights the importance of studying the influence of SDH on quality of life, data on this association in patients with multimorbidity are scarce. Lawson et al. [18] investigated the relationship between multimorbidity and quality of life using a score that combined both physical and mental health function, based on data collected in the Scottish Health Survey. In line with the present findings, those authors observed an association between poorer quality of life and (i) increasing number of comorbid diseases and (ii) greater economic deprivation. In contrast to our findings, they detected no significant influence of sex on quality of life.

Von dem Knesebeck et al. [19] also analysed the association between socioeconomic status and patient-reported outcomes, including HRQoL, in a cohort of patients aged 65–85 with multimorbidity. Among the socioeconomic variables studied, a correlation was found between a lower monthly income and poorer health status, in agreement with the present findings. However, the authors reported no significant correlations between health status and either occupation or educational level.

Park et al. [20] used the EQ-5D-5L scale to measure QoL in older patients, and observed that female sex and lower socioeconomic level were significantly associated with a lower EQ-5D-5L score. Another study of the relationship between educational level and quality of life [21] reported significant associations between worse quality of life and both low educational level and lower monthly income.

Greater feelings of loneliness have been related to poorer quality of life in elderly patients with multimorbidity [22]. Specifically, feeling of loneliness were found to influence the psychophysical health of elderly individuals. While the present study did not include any variables related to social loneliness, we observed a correlation between a lower level of social support and poorer quality of life.

As stated above, few studies have analysed the relationship between multimorbidity, SDH, and quality of life, highlighting the need for greater research in this area to better understanding the underlying contributing factors and the types of interventions required to address multimorbidity. Older patients with low socioeconomic status experience a ‘double burden of disease’. They are at higher risk of multimorbidity and, compared with matched patients with a higher socioeconomic status, are worse off in terms of key outcomes such as functional status, health related quality of life, and self-rated health [19]. In addition, the relationship between socioeconomic factors and quality of life is influenced by psychosocial factors including self-efficacy, coping behaviours, social contacts, social support, and psychosocial stress; behavioural factors such as activity level, smoking, and alcohol consumption; and material factors including living conditions [19]. Although supporting scientific evidence is limited, interventions targeting psychosocial and lifestyle-related factors, applied at the patient level, could help minimize multimorbidity [6]. A clinical trial is currently underway in Ireland to evaluate the effectiveness of social interventions at the primary care level on patients’ health in disadvantaged areas of the country [23].

SDH are undoubtedly key contributors to social inequity, which in turn arise as a consequence of sociopolitical and socioeconomic factors. They also generate vulnerabilities that affect all aspects of the individual’s life, including health [24]. Moreover, synergies between different factors can exponentially increase the risk of multimorbidity, beyond their expected additive effects [25].

One such example is the association between lower quality of life and both socioeconomic level and deprivation index: this relationship is influenced by multiple factors, one of which is the “inverse care law” [26,27]. This law reflects the fact that health service availability does not correspond to the greater needs of disadvantaged populations, resulting in insufficient health resources to manage patients’ conditions. Furthermore, this effect can be exacerbated by the fact that disadvantaged social groups often have reduced personal capacity, community support [18], and access to healthy food [20], factors that could potentially mitigate the impact of multimorbidity.

Knowledge of the influence of social variables on health behaviours, and consideration by healthcare providers of patients’ socioeconomic context, health-related quality of life, and preferences, can help ensure a more effective approach to the management of multimorbidity. Understanding of factors that influence quality of life can enable implementation of strategic measures based on SDH. Consideration of quality of life as perceived by the patient is also important to improve the care of elderly patients with multimorbidity, and to facilitate individualized therapy and increased involvement in self-care.

### Strengths and weaknesses

A key strength of this study is the use of data derived from a multicentre study in which primary care professionals participated. The availability of these data enabled a broader assessment of participating patients, including data relating to a range of psychosocial variables that are frequently omitted from such studies (e.g. quality of life, social support, and urban vulnerability).

Limitations of this study are those intrinsically related to the measurement of social variables. Subjectivity in responses can lead to a higher degree of interpatient variability. Furthermore, it is difficult to separately analyse closely interrelated variables that are ultimately part of larger, multidimensional concepts. To maximise internal validity, we used the version of the DUFSS questionnaire validated for use in non-institutionalized elderly individuals [13], a population that closely corresponds to that of the present study.

Measurement of variables that may affect quality of life is important, as poorer self-perceived quality of life can be associated with poorer adherence to treatment, which in turn is related to greater urban vulnerability and lower functional support [28]. Other independent variables that may be related to quality of life but that were not considered in our analysis include psychosocial factors such as self-efficacy, coping behaviour, social contacts, social support, and psychosocial stress, and behavioural factors such as physical activity, smoking, and alcohol consumption [19].

Lastly, limiting the study to patients aged 65–74 with multimorbidity can result in a loss of external validity if the results are extrapolated to the rest of the population. However, the population aged 65–74 corresponds to an increasingly larger stratum of the population, not only in Spain but in developed countries in general [29]. Furthermore, the large sample size and the fact that this study was conducted via coordinated efforts in 3 different regions in Spain lend greater validity to our findings in terms of potential for extrapolation to the broader population in this age range.

## Conclusion

Our findings showed the association between SDH and quality of life in patients aged 65–74 with multimorbidity and polypharmacy. A greater number of illnesses, female gender, lower educational level, urban vulnerability, and reduced social support were all identified as variables associated with poorer quality of life in this population, underscoring the need for a biopsychosocial approach to the care of these patients. Consideration by healthcare professional of the socioeconomic context, quality of life, and living conditions of these patients can help ensure a more effective approach to the management of multimorbidity.

## Acknowledgments

The authors thank all healthcare professionals and patients from the Primary Healthcare Centres that participated in this study.

## MULTIPAP GROUP

### Lead authors for the MULTIPAP Study group

Alexandra Prados Torres (Aragonese Institute of Health Sciences (IACS), IIS Aragón, Miguel Servet University Hospital, Spain), Juan Daniel Prados Torres (Multiprofessional Teaching Unit for Family and Community Care Primary Care District Málaga-Guadarhorce. Málaga), Isabel del Cura (Research unit. Primary Health Care Management Madrid. Spain).

#### Coordinating Committee

Mercedes Aza-Pascual-Salcedo (EpiChron Research Group, Aragon Health Sciences Institute (IACS), IIS Aragón, RICAPPS ISCIII, Aragon Health Service (SALUD), Zaragoza, Spain), Antonio Gimeno-Miguel (EpiChron Research Group, Aragon Health Sciences Institute (IACS), IIS Aragón, RICAPPS ISCIII, Miguel Servet University Hospital, Zaragoza, Spain), Francisca González Rubio (EpiChron Research Group, Aragon Health Sciences Institute (IACS), IIS Aragón, RICAPPS ISCIII, Zaragoza, Spain), Cristina M Lozano Hernández (Research unit. Primary Health Care Management Madrid. Spain), Juan A. López-Rodríguez (Research unit. Primary Health Care Management Madrid. Spain, RICAPPS ISCIII), Beatriz Poblador-Plou (EpiChron Research Group, Aragon Health Sciences Institute (IACS), IIS Aragón, RICAPPS ISCIII, Miguel Servet University Hospital, Zaragoza, Spain), Francisca Leiva Fernández (Multiprofessional Teaching Unit for Family and Community Care Primary Care District Málaga-Guadarhorce. Málaga, RICAPPS ISCIII), Fernando López-Verde (Las Delicias Health Center (Centro de Salud las Delicias), Málaga-Guadalhorce Health District, Málaga, Spain), Victoria Pico-Soler (EpiChron Research Group, Aragon Health Sciences Institute (IACS), IIS Aragón, RICAPPS ISCIII, Aragon Health Service (SALUD), Zaragoza, Spain), Ma Josefa Bujalance-Zafra (La Victoria Health Center (Centro de Salud la Victoria), Málaga-Guadalhorce Health District, Málaga, Spain), Luis A. Gimeno-Feliu (EpiChron Research Group, Aragon Health Sciences Institute (IACS), IIS Aragón, RICAPPS ISCIII, Aragon Health Service (SALUD), Zaragoza, Spain), Marisa Rogero-Blanco (Instituto de Investigación Sanitaria Gregorio Marañon IISGM, RICAPPS ISCIII, Ricardos General Health Center, Madrid, Spain), Francisca García-de-Blas (Dr Mendiguchia Carriche Health Center, RICAPPS ISCIII Madrid, Spain), Marcos Castillo Jimena (Coín Health Center, Málaga-Guadalhorce Health District, Málaga, Spain;), Marcos Alonso-García (Preventive Medicine Unit, University Hospital Alcorcón Foundation, Madrid, Spain), Alessandra Marengoni (Department of Clinical and Experimental Sciences, University of Brescia, Brescia, Italy), Jesús Martín Fernández (Multiprofessional Teaching Unit for Family and Community Care, Primary Healthcare Centre (PHC) Oeste, Madrid, Spain), Elena Polentinos Castro (Research unit. Primary Health Care Management Madrid. Spain), José María Valderas Martínez (National University of Singapur, RICAPPS ISCIII), María del Pilar Barnestein-Fonseca (CUDECA Foundation, Biomedical Research Institute of Málaga –IBIMA, Malaga University, Spain), Miguel Domínguez-Santaella (PHC Victoria, Malaga-Guadalhorce Health District, Andalusian Health Service, Malaga, Spain), Nuria García-Agua-Soler (Department of Pharmacology, Faculty of Medicine, Malaga University, Spain), María Isabel Márquez-Chamizo (PHC Carranque, Malaga-Guadalhorce Health District, Andalusian Health Service, Málaga, Spain), José María Ruiz-San-Basilio (PHC Coín, Malaga-Guadalhorce Health District, Andalusian Health Service, Coín, Málaga, Spain), José María Abad-Díez (Lozano Blesa Hospital, Zaragoza, Spain), Marta Alcaraz Borrajo (Subdirectorate General of Pharmacy and Health Products), Gloria Ariza Cardiel (Multiprofessional Teaching Unit for Family and Community Care Primary Care PHC Oeste, Madrid, Spain), Amaya Azcoaga Lorenzo (PHC Pintores, Madrid, Spain), Marta Alcaraz Borrajo (Subdirectorate General of Pharmacy and Health Products), Ana Cristina Bandrés-Liso (EpiChron Research Group, Aragon Health Sciences Institute (IACS), IIS Aragón, RICAPPS ISCIII, Aragon Health Service (SALUD), Zaragoza, Spain), Amaia Calderon-Larrañaga (EpiChron Research Group, Aragon Health Sciences Institute (IACS), IIS Aragón, RICAPPS ISCIII, Zaragoza, Spain, Aging Research Center, Department of Neurobiology, Care Sciences and Society, Karolinska Institute, Stockholm University, Stockholm, Sweden) Mercedes Clerencia-Sierra (EpiChron Research Group, Aragon Health Sciences Institute (IACS), IIS Aragón, RICAPPS ISCIII, Miguel Servet University Hospital, Aragon Health Service (SALUD), Zaragoza, Spain), Javier Marta-Moreno (IIS Aragón, Miguel Servet University Hospital, Aragon Health Service (SALUD), Zaragoza, Spain), Christiane Muth (Department of General Practice and Family Medicine, Medical Faculty OWL, University of Bielefeld, Germany), Antonio Poncel-Falcó (EpiChron Research Group, Aragon Health Sciences Institute (IACS), IIS Aragón, RICAPPS ISCIII, Aragon Health Service, Zaragoza, Spain), Ana Isabel González González (Technical Support Unit, Primary Care Management, Madrid Health Service), Virginia Hernández Santiago (Ninewells Hospital & Medical School, Dundee, UK), Angel Mataix SanJuan (Subdirectorate General of Pharmacy and Health Products, Madrid, Spain), Ricardo Rodríguez Barrientos (Research unit. Primary Health Care Management, Madrid, Spain), Mercedes Rumayor Zarzuelo (Public Health Centre Coslada, Área II Subdirectorate of Prevention and Health Promotion, Madrid, Spain), Luis Sánchez Perruca (General System Information Management, Primary Healthcare Management, Madrid Health Service, Spain), Teresa Sanz Cuesta (Research unit. Primary Health Care Management Madrid. Spain), María Eugenia Tello Bernabé (PHC El Naranjo, Madrid, Spain).

#### Clinical Investigators in Primary Healthcare Centres (PHC) MULTIPAP GROUP

**Andalucía, Spain: PHC Alhaurín el Grande** (Alhaurín el Grande): Javier Martín Izquierdo, Macarena Toro Sainz. **PHC Carranque** (Málaga): María José Fernández Jiménez, Esperanza Mora García, José Manuel Navarro Jiménez. **PHC Ciudad Jardín**, (Málaga): Leovigildo Ginel Mendoza, Luz Pilar de la Mota Ybancos, Jaime Sasporte Genafo. **PHC Coín**, (Málaga): María José Alcaide Rodríguez, Elena Barceló Garach, Beatriz Caffarena de Arteaga, María Dolores Gallego Parrilla, Catalina Sánchez Morales. **PHC Delicias**, (Málaga): María del Mar Loubet Chasco, Irene Martínez Ríos, Elena Mateo Delgado. **PHC La Roca**, (Málaga): Esther Martín Aurioles. **PHC Limonar**, (Málaga): Sylvia Hazañas Ruiz. **PHC Palmilla**, (Málaga): María Auxiliadora Nieves Muñoz Escalante. **PHC Puerta Blanca**, (Málaga): Enrique Leonés Salido, María Antonia Máximo Torres, Maria Luisa Moya Rodríguez, María Encarnación Peláez Gálvez, José Manuel Ramírez Torres, Cristóbal Trillo Fernández. **PHC Tiro Pichón**, (Málaga): María Dolores García Martínez Cañavate, María del Mar Gil Mellado, María Victoria Muñoz Pradilla. **PHC Vélez Sur** (Vélez Málaga): María José Clavijo Peña, José Leiva Fernández, Virginia Castillo Romero. **PHC Vera**, (Vera, Almería): Rubén Vázquez Alarcón. **PHC Victoria**, (Málaga): Rafael Ángel Maqueda, Gloria Aycart Valdés, Ana María Fernández Vargas, Irene García, Antonia González Rodríguez, María Carmen Molina Mendaño, Juana Morales Naranjo, Francisco Serrano Guerra.

**Aragón, Spain: PHC Alcorisa**, (Alcorisa, Teruel): Carmen Sánchez Celaya del Pozo. **PHC Delicias Norte**, (Zaragoza): José Ignacio Torrente Garrido, Concepción García Aranda, Marina Pinilla Lafuente, M^a^ Teresa Delgado Marroquín. **PHC Picarral**, (Zaragoza): M^a^ José Gracia Molina, Javier Cuartero Bernal, M^a^ Victoria Asín Martín, Susana García Domínguez. **PHC Fuentes de Ebro**, (Zaragoza): Carlos Bolea Gorbea. **PHC Valdefierro**, (Zaragoza): Antonio Luis Oto Negre. **PHC Actur Norte**, (Zaragoza): Eugenio Galve Royo, M^a^ Begoña Abadía Taira. **PHC Alcañiz**, (Teruel): José Fernando Tomás Gutiérrez. **PHC Sagasta - Ruiseñores**, (Zaragoza): José Porta Quintana, Valentina Martín Miguel, Esther Mateo de las Heras, Carmen Esteban Algora. **PHC Ejea** (Ejea de los Caballeros, Zaragoza): María Teresa Martín Nasarre de Letosa, Elena Gascón del Prim, Noelia Sorinas Delgado, María Rosario Sanjuan Cortés. **PHC Canal Imperial - Venecia**, (Zaragoza): Teodoro Corrales Sánchez. **PHC Canal Imperial - San José Sur**, (Zaragoza): Eustaquio Dendarieta Lucas. **PHC Jaca** (Jaca, Huesca): María del Pilar Mínguez Sorio. **PHC Santo Grial**, (Huesca): Adolfo Cajal Marzal.

**Madrid, Spain: PHC Mendiguchía Carriche** (Leganés): Francisca García De Blas González Eduardo Díaz García, Juan Carlos García Álvarez, Cristina Guisado Pérez, Alberto López García Franco, Maria Elisa Viñuela Benitez. **PHC El Greco** (Getafe): Ana Ballarín González, Maria Isabel Ferrer Zapata, Esther Gómez Suarez, Fernanda Morales Ortiz, Lourdes Carolina Peláez Laguno, José Luis Quintana Gómez, Enrique Revilla Pascual. **PHC Cuzco** (Fuenlabrada): M Ángeles Miguel Abanto. **PHC El Soto** (Móstoles): Blanca Gutiérrez Teira, **PHC General Ricardos** (Madrid): Francisco Ramón Abellán López, Carlos Casado Álvaro, Paulino Cubero González, Santiago Manuel Machín Hamalainen, Raquel Mateo Fernández, Cesar Sánchez Arce. **PHC Ibiza** (Madrid): Jorge Olmedo Galindo. **PHC Las Américas** (Parla): Claudia López Marcos, Soledad Lorenzo Borda, Juan Carlos Moreno Fernández, Belén Muñoz Gómez, Enrique Rodríguez De Mingo. **PHC María Ángeles López** (Leganés): Juan Pedro Calvo Pascual, Margarita Gómez Barroso, Beatriz López Serrano, María Paloma Morso Peláez, Julio Sánchez Salvador, Jeannet Dolores Sánchez Yépez, Ana Sosa Alonso. **PHC M^a^ Jesús Hereza** (Leganés): María del Mar Álvarez Villalba. **PHC Pavones** (Madrid): Purificación Magán Tapia. **PHC Pedro Laín Entralgo** (Alcorcón): María Angélica Fajardo Alcántara, María Canto De Hoyos Alonso, María Aránzazu Murciano Antón. **PHC Pintores** (Parla): Manuel Antonio Alonso Pérez, Ricardo De Felipe Medina, Amaya Nuria López Laguna, Eva Martínez Cid De Rivera, Iliana Serrano Flores, María Jesús Sousa Rodríguez. **PHC Ramón y Cajal** (Alcorcón): María Soledad Núñez Isabel, Jesús M^a^ Redondo Sánchez, Pedro Sánchez Llanos, Lourdes Visedo Campillo.

## Funding

This study was funded by the Carlos III Institute of Health, Ministry of Science and Innovation (Spain) (Grant Numbers PI15/00276, PI15/00572, PI15/00996); the European Regional Development Fund (“A way to build Europe”); the Network for Research on Chronicity, Primary Care, and Health Promotion (RICAPPS) awarded as part of the call for the creation of Health Outcomes-Oriented Cooperative Research Networks (grant number RD21/0016/0019, RD21/0016/0015 and RD21/0016/0027); the Gobierno de Aragón (grant number B01_23R); and co-funded with European Union’s NextGenerationEU funds.

## Competing interests

The authors declare no competing interests.

## Data availability

The Aragon Ethics Committee approved this research without considering the option of data sharing. The data include sensitive clinical information about patients, and there are therefore ethical and legal restrictions to sharing the data set. The data are part of the MULTIPAP study and can be requested by contacting the Aragon Ethics Committee at the email address. Data can also be requested by contacting the Primary Care Management of Madrid at the email address, and the Technical Direction of Teaching and Research at the email address. The MULTIPAP Group may establish future collaborations with other groups based on the same data. Data will be available from the authors upon reasonable request and with permission of the project’s principal investigators (Alexandra Prados-Torres; Daniel Prados-Torres; Isabel del Cura). However, each new project based on these data must be first submitted to CEICA for approval.

## Ethics statement

The trial was designed in accordance with the basic ethical principles of beneficence, autonomy, non-maleficence and justice, and it was conducted in accordance with the rules of Good Clinical Practice outlined in the Oviedo Convention (1997) and the most recent Declaration of Helsinki. Written informed consent of patients was required. Data confidentiality and anonymity was ensured, according to the provisions of Spanish Law 15/1999, both during the implementation phase of the project and in any resulting presentations or publications.

This study, as well as the written model form, was approved by the Clinical Research Ethics Committees of Aragón (CEICA, PI15/0217) and was favourably evaluated by the Research Ethics Committee of the Province of Malaga and by the Central Committee of Primary Care Research of the Community of Madrid.

## Notes

### Competing Interest Statement

The authors have declared no competing interest.

### Funding Statement

Yes

### Author Declarations

This study was approved by the Clinical Research Ethics Committees of Aragón (CEICA, PI15/0217) and was favourably evaluated by the Research Ethics Committee of the Province of Malaga and by the Central Committee of Primary Care Research of the Community of Madrid. The recruiting physicians provided the patient with detail information about the study, they confirmed the patient's eligibility and obtained the patient's written informed consent.

